# Performance of an artificial intelligence-based smartphone app for guided reading of SARS-CoV-2 lateral-flow immunoassays

**DOI:** 10.1101/2022.02.16.22271042

**Authors:** David Bermejo-Peláez, Daniel Marcos-Mencía, Elisa Álamo, Nuria Pérez-Panizo, Adriana Mousa, Elena Dacal, Lin Lin, Alexander Vladimirov, Daniel Cuadrado, Jesús Mateos-Nozal, Juan Carlos Galán, Beatriz Romero-Hernández, Rafael Cantón, Miguel Luengo-Oroz, Mario Rodríguez-Domínguez

## Abstract

**Objectives:** To evaluate an artificial intelligence-based smartphone application to automatically and objectively read rapid diagnostic test (RDT) results and assess its impact on COVID-19 pandemic management.

**Methods:** Overall, 252 human sera from individuals with PCR-positive SARS-CoV-2 infection were used to inoculate a total of 1165 RDTs for training and validation purposes. We then conducted two field studies to assess the performance on real-world scenarios by testing 172 antibody RDTs at two nursing homes and 92 antigen RDTs at one hospital emergency department.

**Results:** Field studies demonstrated high levels of sensitivity (100%) and specificity (94.4%, CI 92.8-96.1%) for reading IgG band of COVID-19 antibodies RDTs compared to visual readings from health workers. Sensitivity of detecting IgM test bands was 100% and specificity was 95.8%, CI 94.3-97.3%. All COVID-19 antigen RDTs were correctly read by the app.

**Conclusions:** The proposed reading system is automatic, reducing variability and uncertainty associated with RDTs interpretation and can be used to read different RDTs brands. The platform can serve as a real time epidemiological tracking tool and facilitate reporting of positive RDTs to relevant health authorities.

## Introduction

To control COVID-19 pandemic, timely and accurate early-detection strategies of SARS-CoV-2 infections have been critical to slow down the spread of the virus. The use of rapid diagnostic tests (RDTs), both for detection of antibodies and antigens, has contributed to improve COVID-19 testing capacity reducing costs of diagnosis and allowing for fastest results (1). First, COVID-19 RDTs were intended to be used just by professional healthworkers, who have extensive experience in the use of this tool for different infectious diseases (2, 3). Later, multiple Health Ministries have approved home testing kits improving the accessibility to testing and taking pressure off from health institutions. Nevertheless, self-testing strategies have some limitations, the general population is not familiar with the use of RDTs and a minimum training is needed for sampling, testing and result interpretation. Furthermore, as it has been seen during the latest Omicron wave (4), many results go unreported, impairing post-testing counseling and epidemiological surveillance.

Combining RDTs with digital tools, artificial intelligence (AI) and mobile health approaches can help standardize result interpretation and facilitate immediate reporting and monitoring of results (5). The use of AI to automatically interpret photographs of RDTs has been also recently proposed (6, 7). Here, we describe the development and field validation of a mobile-based tool for reading and reporting multiple types of SARS-COV-2 RDTs which is connected to a real-time epidemiological monitoring web platform.

## Methods

This study was divided into two phases. First, the training and validation of an AI algorithm for the automatic interpretation of RDTs. Second, two field studies to assess the performance of the AI-based system for reading both COVID-19 antibodies and antigen RDTs in real-world scenarios. Ethics approval for the study was obtained from the Clinical Research Ethics Committee of the Ramón y Cajal University Hospital (Ref. 127/21).

### Algorithm Training and validation dataset

For generating the training image dataset, twelve human sera from SARS-CoV-2 positive PCR patients with a positive ELISA test (Vircell Spain S.L.U., Granada, Spain) were used. Each serum sample was serially diluted until it reached a negative result when inoculated in a COVID-Ab test. Each dilution was tested in three replicates for each of the three brands tested (2019-nCoV IgG/IgM Rapid Test Cassette (Hangzhou AllTest Biotech Co., Ltd.); Panbio COVID-19 IgG/IgM Rapid Test Device (Abbott); UNscience COVID-19 IgG/IgM Rapid Test (Wuhan UNscience Biotechnology Co., Ltd.), resulting in 433 RDTs inoculated (61 IgG+/IgM+; 166 IgG+/IgM-; 43 IgG-/IgM+; 164 negative). Additionally, twelve COVID-Ag RDTs (Panbio COVID-19 Ag Rapid Test Device (Abbott) (6+,6-) were also included to train the algorithm to read not only three-band tests (such as the COVID-19 antibody tests used in this study) but also two-band RDTs such as COVID-19 antigen tests. The entire training dataset consisted of 3614 images.

For collecting the independent validation dataset, 240 human sera samples independent from the ones used for training were used to inoculate 720 COVID-Ab RDTs (each serum was tested in triplicate using the aforementioned brands). The samples were selected ensuring all possible results are well represented along the dataset (108 IgG+/IgM+; 321 IgG+/IgM-; 27 IgG-/IgM+; 264 negative).

Each RDT was visually read by multiple observers (from 3 to 5) and the ground truth was established as the majority result from the total of analyzers. Each inoculated RDT was digitized by using the TiraSpot mobile app (Spotlab, Madrid, Spain) for guided and standardized acquisition ensuring correct positioning of RDTs in the image and using a total of 9 smartphone models. Results data was uploaded to a web platform.

### Field validation studies

The workflow for the field studies was as follows: a health professional digitized the RDTs by using the app, was asked for recording the visual interpretation of the test result, images were uploaded to the telemedicine platform and processed by the AI algorithm, and discrepancies between the interpretation made by the health professional and that obtained by the algorithm were subsequently reviewed by an external health professional through the platform.

The first field study used the system as part of a seroprevalence study conducted in two nursing homes in Madrid (Spain). A total of 172 vaccinated health care personnel were included in this study from which a finger-prick blood sample was taken and inoculated into SARS-CoV-2 Rapid Antibody Test (Roche). A trained nurse digitized the RDTs and record their result using the application.

The second field validation field study tested the system to read also COVID-19 antigen tests composed of two bands (Panbio COVID-19 Ag Rapid Test Device, Abbot). This study was carried out at the Emergency Department of the Ramón y Cajal Hospital (Madrid, Spain), where 92 individuals’ nasal swabs were inoculated in antigen tests, and digitized by experienced health professionals using the app.

All images were acquired in very diverse real-world conditions (including different environmental illuminations and shades).

## Results

### AI algorithm training and app validation

All images acquired with the app were uploaded to a cloud platform where the AI algorithm processed the photographs to predict its result interpretation. As shown in **Table 1A**, when comparing the visual interpretations (used as ground truth) against the AI algorithm, the performance was high for all brands of RDTs tested, obtaining a mean sensitivity and specificity of 98% and 100% respectively for detecting the IgG band and a mean sensitivity and specificity of 80% and 89% for the detection of the IgM band.

**Table 1.** Performance of the AI algorithm for predicting RDTs results with respect to human visual reading in the validation set (**A**), in the field study for reading antibodies RDTs (Ab) (**B**) and in the field study when reading antigen RDTs (Ag) (**C**).

| **A** | Model (Ab) | Band | AUC [95% CI] | SN [95% CI] | SP [95% CI] | N |
| --- | --- | --- | --- | --- | --- | --- |
|  | Abbott | IgG | 99.5 [98.7,100] | 96.4 [94.1,98.8] | 100 [100,100] | 94-, 145+ |
|  |  | IgM | 92.5 [85.4,99.6] | 80.8 [75.8,85.8] | 90.7 [87.0,94.3] | 184-, 55+ |
|  | UNScience | IgG | 100 [100,100] | 100 [100,100] | 100 [100,100] | 100-, 140+ |
|  |  | IgM | 89.5 [83.7,95.2] | 80.0 [74.9,85.1] | 88.6 [84.6,92.6] | 214-, 26+ |
|  | AllTest | IgG | 99.8 [99.4,1] | 97.9 [96.1,99.7] | 100 [100,100] | 96-, 144+ |
|  |  | IgM | 90.6 [85.0,96.1] | 79.6 [74.5,84.7] | 86.0 [81.6,90.4] | 186-, 54+ |
|  | Global | IgG | 99.8 [99.5,100] | 98.1 [97.1,99.1] | 100 [100,100] | 290-, 429+ |
|  |  | IgM | 90.8 [87.4,94.3] | 80.0 [77.1,82.9] | 89.0 [86.2,90.9] | 584-, 135+ |
| **B** | Model (Ab) | Band | AUC [95% CI] | SN [95% CI] | SP [95% CI] | N |
|  | Roche | IgG | 100 [100,100] | 100 [100,100] | 94.4 [92.8,96.1] | 18-, 154+ |
|  |  | IgM | 99.6 [96.0,100] | 100 [100,100] | 95.8 [94.3,97.3] | 166-, 6+ |
| **C** | Model (Ag) | Band | AUC [95% CI] | SN [95% CI] | SP [95% CI] | N |
|  | Abbott | NA | 100 [100,100] | 100 [100,100] | 100 [100,100] | 58-, 30+ |

### Validation in real-world scenarios

From the 172 RDTs used in this study (5 IgG+/IgM+; 149 IgG+/IgM-; 1 IgG-/IgM+; 17 negative), we only found 9 discrepancies between test result interpretation made by the health professional and the AI algorithm. From these 9 cases, two of them were incorrectly classified by the algorithm due to an incorrect image acquisition with the app. The remaining discrepant cases were further reviewed by a second professional, and the AI-based system allowed to detect and modify the result with respect to the initial healthworker interpretation in 4 cases by confirming the result predicted by the algorithm.

The overall performance of the algorithm with respect to the ground truth is shown in **Table 1B**. It should be noted that the performance of the system is high even when used with a RDT different from those used for training the algorithm, suggesting its potential use with any RDT on the market. The slight disparity in the performance of IgM band identification in antibody RDTs between the validation set and this field study may be explained by the presence of very faint signals which were almost invisible in the photographs.

Regarding the second field study for reading COVID-19 antigen RDTs, we found that all tests used and digitized using the TiraSpot app (58 negative, 30 positive) were correctly interpreted by the proposed system (**Table 1C**), demonstrating that the system can also be applied for reading two-band (control and test) as well as three-band tests (IgG, IgM and control).

## Discussion

We describe the usefulness of an app for reading and result interpretation of lateral-flow RDTs for SARS-CoV-2 testing. Additionally, results are sent to a telemedicine platform which allows for case identification and confirmation, quality control and real-time monitoring.

Our AI algorithm demonstrates excellent performance, especially in prospective validation in real-life scenarios and for both antibodies and antigen detection tests. The algorithm performed as well in RDTs brands which were not used at all for training purposes, making the solution suitable for other RDTs, including other diseases. Compared with previous studies (6, 7), our system is able to identify individual bands of the RDTs allowing complex results reading and sending them in real-time to a cloud platform. A requirement and limitation of the proposed system is the correct acquisition of the image (acquisition error in the field studies <0.8%).

In conclusion, the use of TiraSpot (**Figure 1**) is a useful tool for reporting, real-time monitoring and quality control, as the results can be reviewed by specialists when needed. This is especially important in contexts where massive testing is to be done and the likelihood of subjectivity and errors in the interpretation of the result is higher. It is also important in the validation of self-diagnostic tests performed by untrained users, as it avoids the loss of information in case it is not notified by the user and provides an efficient system to confirm and report data, which has been a key challenge during the Omicron wave (4).

**Figure 1.**
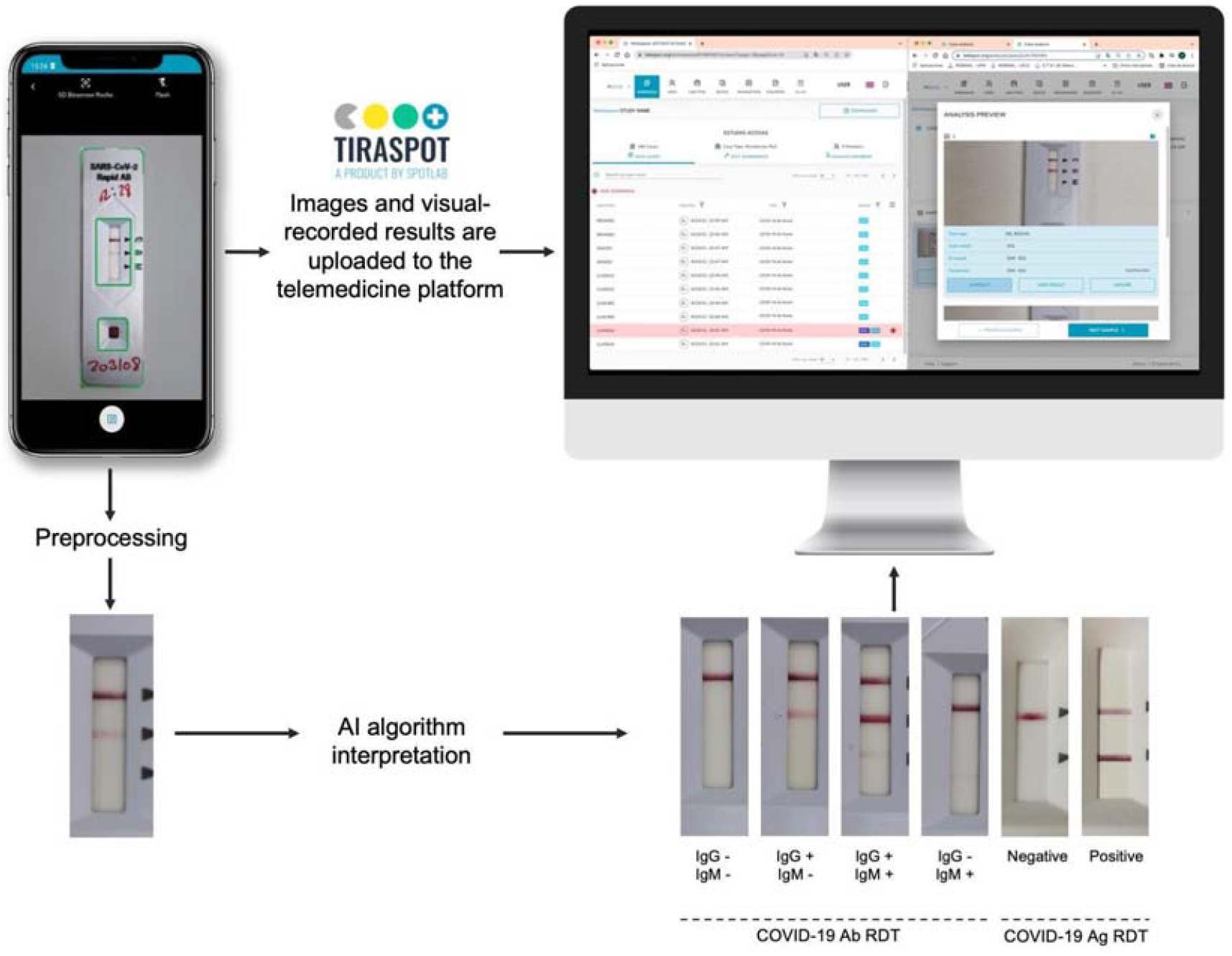
Tiraspot system is composed of (1) a mobile app for test digitization and result recording, (2) an AI model for RDT result interpretation and (3) a web platform where all collected data can be visualized and allows for result corrections in the cases in which a discrepancy exists between AI and user interpretation.

## Data Availability

Data produced in the present study will be available upon reasonable request to the authors

